# The higher benefit of lecanemab in males compared to females in CLARITY AD is probably due to a real sex effect

**DOI:** 10.1101/2024.07.11.24310278

**Authors:** Daniel Andrews, Simon Ducharme, Howard Chertkow, Maria Pia Sormani, D. Louis Collins, the Alzheimer’s Disease Neuroimaging Initiative

## Abstract

**INTRODUCTION:** The Phase 3 trial CLARITY AD found that lecanemab slowed cognitive decline by a statistically significant 27% vs. placebo. However, the subgroup analysis indicated a significant sex difference in the effect, and recent work has implied that lecanemab has either no or limited effectiveness in females. To resolve this ambiguity, we used simulations constrained by the trial design to determine whether the difference could be explained by known sex differences in Alzheimer’s progression, or as an isolated random event.

**METHODS:** Simulations were generated using linear mixed models of cognitive decline fit to data from ADNI participants who satisfied CLARITY AD inclusion criteria.

**RESULTS:** The statistically nonsignificant 7.9% sex difference in cognitive decline rate observed in our selected ADNI participants does not explain the trial’s 31% sex difference in lecanemab’s effect. A ≥31% difference occurred randomly in only 12 of our 10,000 simulations, signifying a probability of 0.0012.

**DISCUSSION:** Our results are consistent with those from CLARITY AD. Lecanemab likely affects females and males differently, but we cannot conclude that the drug is ineffective in females.

## 1. Introduction

Alzheimer’s disease (AD) affects males and females at different rates.^1^ Two thirds of patients are female, and lifetime risk in females is twice that of males.^2–4^ Sex differences in genetics, hormones, and other biological mechanisms are hypothesized to contribute to sex differences in the disease progression and, possibly, drug efficacy.^2^ Recent work has therefore called for clinical trials powered to include primary endpoints disaggregated by sex.^2,5^ While no such trial has yet been done in AD, trials of new drugs do often run sex subgroup analyses.^6–8^

Note that the purpose of such analyses is to generate hypotheses for future research, not to evaluate efficacy within each subgroup.^9^ Ideally, statistical interaction tests are used to determine whether one subgroup benefited more from a drug than a corresponding subgroup (e.g., males vs. females).^9^ In some trials, however, subgroup results are only reported visually on a forest plot, where the trial’s primary endpoint and confidence interval are presented for each subgroup individually.^6,7,10^ This format implies to the reader that subgroup-specific effectiveness is being evaluated, despite typical subgroup sizes not being powered to test the trial’s primary hypothesis.^6–9^ Biostatisticians recommend that primary hypothesis tests not be run and reported separately for low-powered subgroups.^9–11^

These complications make forest plots hard to interpret clinically without strong statistical expertise, especially if there are striking differences in results between corresponding subgroups. That exact scenario may have contributed to important misinterpretations of the sex results in CLARITY AD, the Phase 3 trial of the amyloid-targeting drug lecanemab.^7^

CLARITY AD’s primary analysis showed a statistically significant 27% slowing of cognitive decline for lecanemab-treated vs. placebo participants.^7^ This outcome supported the United States Food and Drug Administration (FDA) approval of lecanemab in July 2023.^12–14^

However, a supplementary forest plot from the CLARITY AD results paper (Fig. S1-B in its appendix) indicated a statistically significant sex difference in lecanemab’s primary clinical effect.^7^ Males showed a significant 43% mean slowing of cognitive decline, while females showed a nonsignificant 12% mean slowing.

At least four recent works and a letter to the editor have interpreted those results as suggesting lecanemab might not slow decline in females.^15–19^ Others have focused on the apparent sex difference in effectiveness.^2,5^ The CLARITY AD paper’s main text itself did not explicitly discuss the sex analysis results.^7^ The authors ostensibly concluded that there was no significant sex difference, as has other another recent work, possibly because sex was not significant as a covariate in a subgroup analysis model.^19,20^ Also, the confidence intervals of the male and female subgroups in the results paper’s Fig. S1-B forest plot overlapped the reported 27% cohort mean.^7^

These interpretations could influence clinical decisions on whether lecanemab is prescribed to female patients and might raise concerns about the drug’s indication in both sexes. The question remains: How should clinicians interpret the published CLARITY AD sex results?

Critically, CLARITY AD was not powered to evaluate efficacy separately for each sex.^7^ A too-small sample size could explain the statistical nonsignificance of the effect in the female subgroup, an effect that nonetheless trended toward favoring lecanemab.^7,21^ These facts undermine the conclusion in some papers that lecanemab is ineffective in females.^15–19^

The low power issue does not address lecanemab’s lower clinical effect on females than males, which might be explained exclusively by AD-related sex differences in cognitive decline. For instance, females with prodromal AD have been shown to decline faster than corresponding males.^22–24^ Research has shown that females with AD brain pathology similar to males are more likely to receive clinical AD diagnoses.^25^

On the other hand, CLARITY AD’s sex difference could be a totally random occurrence – a fluke – linked to randomization and participant heterogeneity in disease progression. Longitudinal studies have shown that patients with similar AD severities at baseline can have very different cognitive decline rates.^26–29^ Computer-generated simulations have shown that randomization in AD trials can produce an imbalance of fast- and slow-progressing patients between treatment groups, thus affecting observed effect sizes for drug efficacy.^30^

In this paper, we use sex-specific models of cognitive decline and simulations constrained by the CLARITY AD trial design parameters (e.g., cohort composition, visit scheme, sample size, dropout rate) to test the hypothesis that the trial’s sex difference could be explained by either of the two phenomena above. If both are unlikely, then lecanemab could have affected CLARITY AD females differently than the males.

An expert reading of the trial’s published supplementary forest plot described above will already conclude that the sex difference was unlikely to have been a fluke, since the non-overlap of the confidence limits would suggest a significant interaction test. However, that plot did not report a formal interaction test and does not provide information on whether natural AD sex differences explain the result. By using simulation to empirically evaluate both possibilities, we aim to help the reader conclusively interpret the real trial’s sex subgroup results.

## 2. Methods

### 2.1. Experimental reasoning

To test our hypothesis, we ask two basic questions: (1) Could the CLARITY AD sex difference be explained by known pre-existing sex differences in cognitive decline? (2) Could the sex difference be explained as a fluke difference between subgroups?

One question is answered in each of two experiments. If the answer to either question is “yes,” then our original hypothesis is likely true, implying there was likely no genuine sex difference in lecanemab’s clinical effect in CLARITY AD. If the answer to both questions is “no,” then lecanemab may have had a different effect on the trial’s males and females.

### 2.2. Alzheimer’s Disease Neuroimaging Initiative

Our analyses used participant data from the Alzheimer’s Disease Neuroimaging Initiative (ADNI) database (https://adni.loni.usc.edu), specifically from the ADNI-1, ADNI-GO, ADNI-2, and ADNI-3 cohorts.^31–34^ The ADNI was launched in 2003 as a public-private partnership, led by Principal Investigator Michael W. Weiner, MD. The primary goal of ADNI has been to test whether serial magnetic resonance imaging (MRI), positron emission tomography (PET), other biological markers, and clinical and neuropsychological assessment can be combined to measure the progression of MCI and early AD. For up-to-date information, see www.adni-info.org. ADNI received ethics approval from all participating institutions and received informed consent from all participants.

### 2.3. Simulation cohort participants

We selected ADNI participants who satisfied CLARITY AD inclusion criteria listed on ClinicalTrials.gov.^35^ Data from the selected participants were used in our modeling and simulation experiments. These participants comprise our “simulation cohort.” At baseline, each participant met the clinical criteria for intermediate likelihood AD-related MCI or probable mild AD dementia, were between 50 and 90 years old, and had Mini Mental State Examination (MMSE) scores ≥22 and ≤30. Participants with MCI had global Clinical Dementia Rating (CDR) scores of 0.5 and CDR Memory Box scores ≥0.5. Mild AD dementia participants had global CDR scores of 0.5 or 1.0 and CDR Memory Box scores ≥0.5. All participants had a body mass index >17 and <35 at baseline. Participants were apolipoprotein E4 (APOE4) genotyped and were positive for abnormal amyloid levels at baseline. Amyloid status was determined using ADNI’s reported baseline PET tracer measures of brain amyloid levels (thresholds: PiB SUVR > 1.2 or AV45 SUVR > 1.11),^36,37^ or baseline cerebrospinal fluid measures of amyloid-beta 42 (thresholds: Aβ42 ≤ 980 pg/ml).^38^ The left panel in Fig. 1 shows how the simulation cohort is generated.

**Figure 1.**
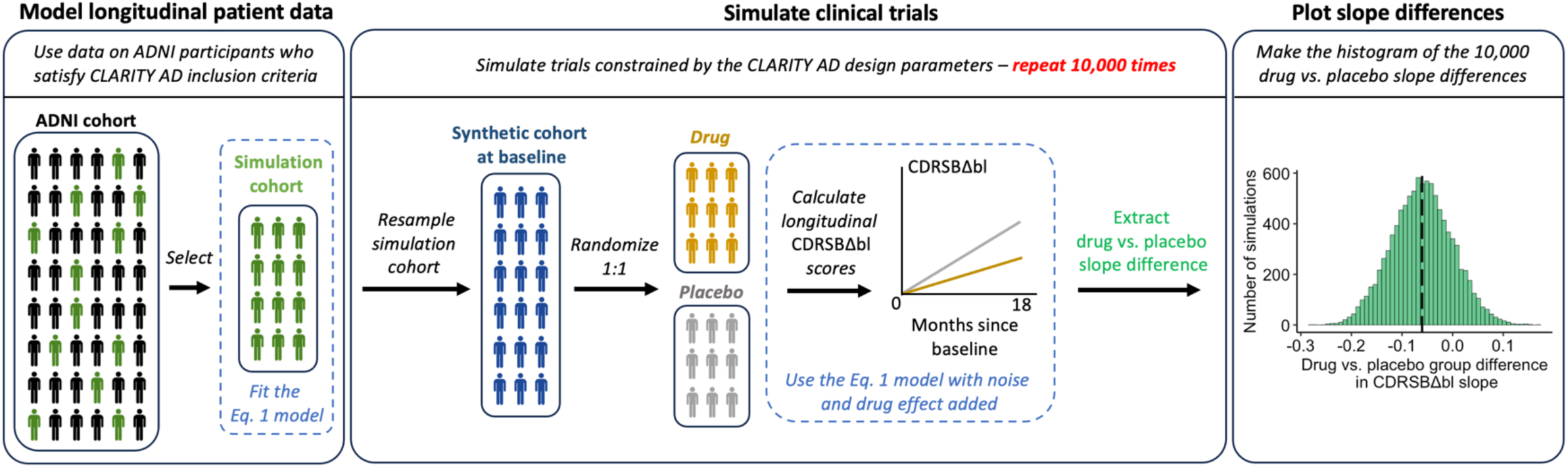
Procedure for generating 10,000 clinical trial simulations constrained by the CLARITY AD design. First, ADNI participants who fit CLARITY AD inclusion criteria are selected to form our “simulation cohort.” We then fit the Eq. 1 model to data from subsets of that cohort in Experiment 1 (only females and only males) and from the full cohort in Experiment 2. The model longitudinally tracks cohort- and participant-level CDRSB change since baseline (CDRSBΔbl) scores. Next, to simulate a trial, we first randomly resample participants from our simulation cohort then add noise to the baseline CDRSB score of each sampled participant. This process generates a cohort of synthetic participants – our “synthetic cohort” – for our simulated trial, for example where the sample size is equal to that specified in the CLARITY AD design. We then randomize those participants to “drug” and “placebo” groups. The synthetic cohort data are then input to the Eq. 1 model that was originally fit to the simulation cohort. With noise added to the model parameters, Eq. 1 is used to calculate CDRSBΔbl scores for each participant at a number of visits that matches the observation scheme of CLARITY AD, up to 18 months. A drug effect is simultaneously injected as a reduction in CDRSBΔbl slope for drug-treated participants relative to placebo. Once the longitudinal CDRSBΔbl scores are generated (i.e., the trial data simulation is complete), the drug vs. placebo group difference in CDRSBΔbl slope is extracted. The extracted slope difference is the observed drug effect size in the simulated trial. This process is repeated 10,000 times. Each simulated trial will have a different observed effect size because of the heterogeneity in natural cognitive decline trajectories in the trial’s unique synthetic cohort. Finally, the observed effect sizes across the 10,000 simulations are plotted in a histogram, where the mean observed drug vs. placebo difference in slope is here indicated by the black dashed line.

### 2.4. Experiment 1

In Experiment 1 we asked: Given the CLARITY AD trial design and cohort, could the observed sex difference in lecanemab’s clinical effect be explained by lecanemab-independent sex differences in cognitive decline trajectories? We answer this question in two parts.

In Part 1 we use a linear mixed effects model to track cognitive decline separately in the male and female participants in our selected ADNI cohort. Our goal is to determine whether there is an inherent average sex difference in cognitive decline rate.

In Part 2 we use the sex-specific models from Part 1 to simulate trials adhering to CLARITY AD design parameters in the hypothetical scenarios of including only males or only females. In these scenarios, a simulated drug slows cognitive decline by 27% vs. placebo (the overall lecanemab effect in CLARITY AD).^7^ The sex-specific placebo decline rate is defined by the estimated slope of the corresponding model from Part 1. If the placebo group’s decline rate is sufficiently different between males and females, a sex difference in the observed drug effect size could result. Our goal here is to determine whether drug effect sizes observed in simulated female-only trials would be different, on average, than effect sizes observed in simulated male-only trials, despite there being no sex difference the drug’s percentage slowing of decline.

A sufficiently large sex difference in cognitive decline trajectories in Part 1 or in possible observed drug effect sizes in Part 2 might explain the CLARITY AD sex difference result.

#### 2.4.1. Part 1: Sex-specific modeling procedure

We fitted a continuous-time linear mixed effects model (Eq. 1) separately to the male and female subsets of our ADNI simulation cohort (see the middle panel of Fig. 1). These models longitudinally track group- and participant-level trajectories in Clinical Dementia Rating Sum-of-Boxes change since baseline (CDRSBΔbl). Increasing CDRSB scores indicate worsening impairment.^39^

All our analyses were done in *R* version 4.3.2 in macOS 14.2.1.^40^ To fit the model, we used the function *lmer* in package *lme4*.^41^ The model is a continuous-time analogue of the categorical-time primary analysis model from CLARITY AD.^7^

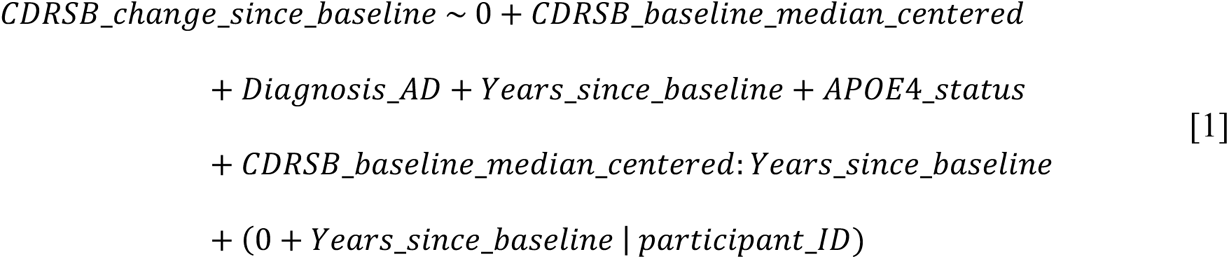

The predictor variables are: median-centered baseline CDRSB scores (*CDRSB*_*baseline*_*median*_*centered*), number of years elapsed since a participant’s baseline visit in ADNI (*Years*_*since*_*baseline*, the continuous time variable), an interaction term for median-centered baseline CDRSB scores with years since baseline (*CDRSB*_*baseline*_*median*_*centered*: *Years*_*since*_*baseline*), AD diagnosis (*Diagnosis*_*AD*, AD = 1 or MCI = 0), and APOE4 status (*APOE*4_*status*, homozygote or heterozygote = 1 or non-carrier = 0). Random slopes for “*Years*_*since*_*baseline*” are encoded by participant. No intercepts are encoded because all participants’ CDRSBΔbl (*CDRSB*_*change*_*since*_*baseline*) scores at baseline are zero.

To complete Part 1 of Experiment 1, we compared the male and female models’ estimated coefficients on “*Years*_*since*_*baseline*” term and their standard errors to evaluate sex difference in cognitive decline rate.

For Part 2, we used the sex-specific models separately to generate synthetic trial data (i.e., synthetic *CDRSB*_*change*_*since*_*baseline* scores over time) in male-only and female-only simulated trials. This process is illustrated in the middle and right panels of Fig. 1 and is described below.

#### 2.4.2. Part 2: Sex-specific trial simulation procedure

Using our female-only trial simulation as the example here, we first randomly sampled female participants from our simulation cohort, with replacement. This resampling procedure generated baseline data for 1,800 synthetic female participants, i.e., the approximate full sample size targeted in the CLARITY AD design. The resampling preserved the approximate baseline AD stage distribution of the real trial, where approximately 60% of participants had MCI and 40% had mild AD dementia at baseline.^7^ For each sampled participant, random noise corresponding to measurement error was added to their baseline CDRSB score.^42^ Noise was also added to the cohort-level and participant-specific model parameters to account for uncertainty and to further differentiate synthetic participants in our resulting “synthetic cohort.”

Next, we randomized these participants 1:1 to “drug” and “placebo” groups. We then used the participants’ baseline data and the estimated parameters from our female-only model to calculate CDRSBΔbl scores (i.e., *CDRSB*_*change*_*since*_*baseline* values) for each participant at 7 visits up to 18 months follow-up. Random noise was added to each participant’s calculated CDRSBΔbl score at each visit to incorporate uncertainty and measurement error. Noise values were drawn from a zero mean normal distribution with standard deviation equal to that of the observation-level residuals in our female-only simulation cohort model. Calculated CDRSBΔbl scores were then rounded to correspond to standard CDRSB 0.5 increments (e.g., 1.1 rounded to 1.0, and 1.4 rounded to 1.5).^39^ Visits were defined to occur every 3 months, and each participant’s specific visit time values had random noise added to simulate an approximate ±1-month window around each visit. This observation scheme approximates that of CLARITY AD.

A drug effect was simultaneously injected as a 27% reduction in the linear slope of drug-treated synthetic participants’ CDRSBΔbl trajectories compared to the placebo participants. The slope for the placebo group is equal to the *β* (i.e., the coefficient) of the “*Years*_*since*_*baseline*” term in the Eq. 1 model fit to our female-only simulation cohort. In this way, our female-only trial simulation model encodes a female-specific rate of cognitive decline for the placebo group, and thus a female-specific decline rate for the drug group. We also encoded 20% participant dropout by the trial’s final observation, as was assumed in the real CLARITY AD sample size calculation.^7^

This overall procedure generates a simulated trial constrained by the CLARITY AD parameters specified in its analysis plan, as described in the main results paper.^7^ The process was repeated 10,000 times to simulate 10,000 female-only trials matching the CLARITY AD design parameters. Each simulation had the same cohort-level trend but a different sampling of participant-level trajectories, thus accounting for randomization and inter-participant heterogeneity in cognitive decline within and across simulations.^30^ Next, we extracted the drug vs. placebo CDRSBΔbl linear slope difference (i.e., the observed drug effect size) from each simulation. This methodology is conceptually analogous to simultaneously running 10,000 identically designed female-only trials in the real world.

We also applied this procedure using the male-only data and model to generate 10,000 male-only trial simulations. Overall, we obtain a set of 10,000 of observed effect sizes for the female-only simulated trials, and 10,000 for the male-only trials.

To finish Experiment 1 Part 2, we evaluated differences between our female and male effect size distributions using a *t*-test to compare means and a Bartlett test to compare variances.^43,44^ We also calculated the percent difference in those means.

While the 10,000 female-only and 10,000 male-only simulated trials all had the same cohort-level drug effect, we hypothesize that the mean and variance of the effect size distributions would be different between the sexes. A difference in means could result because the placebo group’s cognitive decline rate (parameterized as the *β* for the “*Years*_*since*_*baseline*” term from Eq. 1) differed between female-only and male-only simulations. A difference in variance could result because the female-only and male-only cohorts contained different cognitive decline trajectories. Other parameters from Eq. 1 (e.g., the estimated *β* values and standard errors) also differed between the female-only and male-only models and trial simulators.

### 2.5. Experiment 2

In Experiment 2 we asked: Could CLARITY AD’s observed sex difference have been a fluke difference between the male and female subgroups? A fluke here represents an observed sex difference in drug effect when there is no genuine sex difference in the effect (i.e., a Type 1 error).

We answer this question using simulations, posing it technically as: What is the probability of observing a ≥31% difference in a drug’s effect between two equally sized subgroups containing participants selected totally at random? This 31% was the difference between the male and female effects from lecanemab in CLARITY AD. If the probability is sufficiently high (e.g., > 0.05), then the sex difference in CLARITY AD might have been a fluke.

#### 2.5.1. Trial simulation procedure

Here we simulate trials using the same CLARITY AD design parameters as in Experiment 1. We use the same model as in Eq. 1, but here we fit the model to data from all participants in our ADNI simulation cohort. The same trial simulation procedure as in Experiment 1 is then applied, but here using the pooled male and female data. However, this time we randomize 1,800 synthetic participants 1:1 to “Subgroup 1” and “Subgroup 2,” signifying two subgroups of 900 participants selected totally at random with no consideration of sex. This subgroup sample size is approximately equal to that of the male and female subgroups in the CLARITY AD design.^7^

After running 10,000 simulations, we extracted the Subgroup 1 vs. Subgroup 2 CDRSBΔbl linear slope difference (i.e., the observed difference in drug effect between the subgroups) from each simulation. Next, we calculated the proportion of simulations with a between-subgroup difference ≥31%, yielding the probability of at least that difference arising as a fluke.

## 3. Results

### 3.1. Participant demographics

Of the 2,420 ADNI participants available, 644 met the specified CLARITY AD inclusion criteria, with 264 (41%) being female and 380 (59%) being male. The mean baseline ages (± standard deviations) for females and males, respectively, were 71.6 ± 7.3 years and 73.9 ± 6.8 years. The mean number of years of education was 15.3 ± 2.7 for females and 16.4 ± 2.8 for males. 183 females (69%) and 250 males (66%) were APOE4 heterozygotic or homozygotic. At baseline, 76 females (29%) and 102 males (27%) were diagnosed mild AD dementia, with the remaining participants having a baseline diagnosis of AD-related MCI. See Table 1 for the demographic summary.

**Table 1.** Demographics of our simulation cohort participants selected from ADNI.

|  | **Females (n = 264)** | **Males (n = 380)** |
| --- | --- | --- |
| Age (mean years ± std. deviation) | $71.6 \pm 7.3$ | $73.9 \pm 6.8$ |
| Education (mean years ± std. deviation) | $15.3 \pm 2.7$ | $16.4 \pm 2.8$ |
| APOE4 positive | 183 (69%) | 250 (66%) |
| Mild AD dementia | 76 (29%) | 102 (27%) |
| Mild cognitive impairment | 188 (71%) | 278 (73%) |

### 3.2. Experiment 1 results

#### 3.2.1. Part 1: Sex-specific modeling results

Females and males who fit CLARITY AD inclusion criteria had similar CDRSBΔbl trajectories estimated by the Eq. 1 model. We used the Nakagawa conditional *R*^2^ for mixed models as an absolute value goodness-of-fit measure, yielding 0.931 and 0.928 for the male-only and female-only models, respectively, indicating well-fitting models.^45,46^ Key parameters for the male-only and female-only models are summarized in Table 2-A and B, respectively. Fig. 2-A shows estimated cognitive decline trends by sex, as well as real trajectories for individual participants. There was no statistically significant sex difference in estimated cognitive decline rate between the male and female models, as the confidence intervals on the “*Years*_*since*_*baseline*” *β* value of each sex overlaps the opposing sex *β* value on this term (see Table 2-A and B).

**Figure 2.**
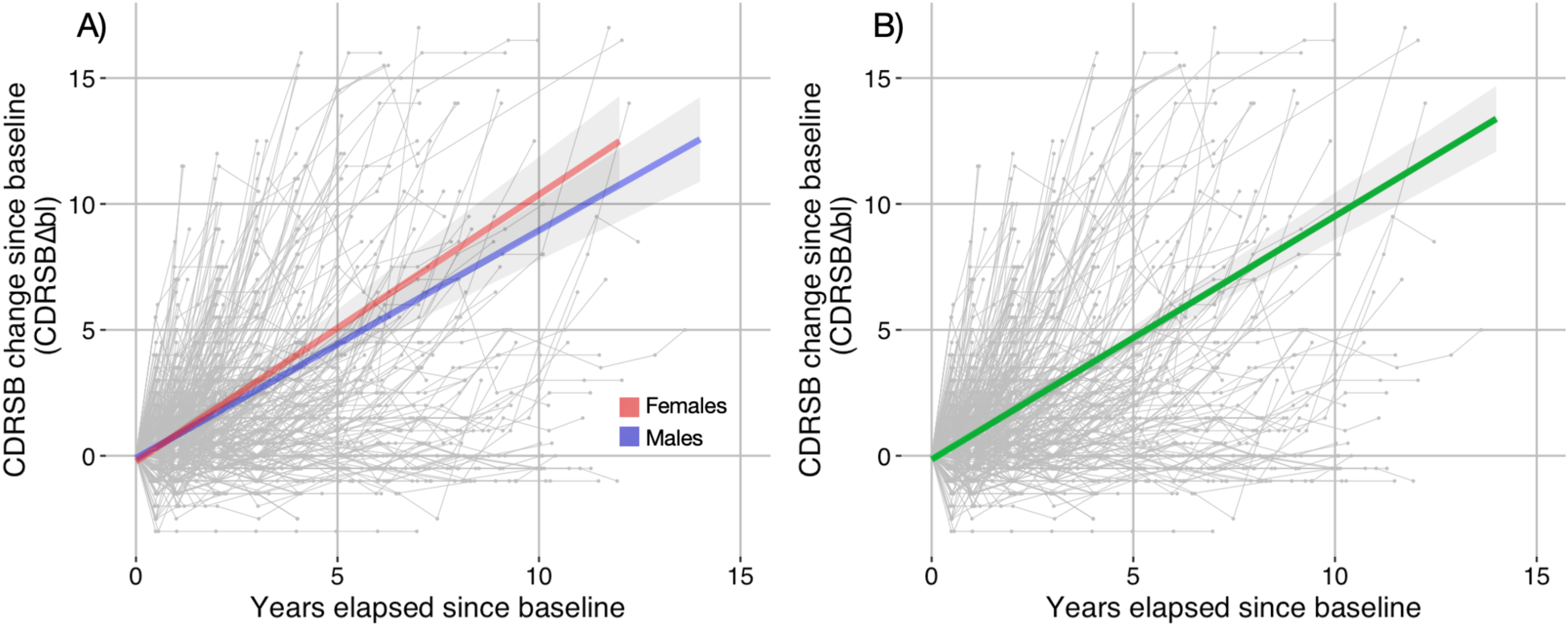
CDRSB change since baseline (CDRSBΔbl) trajectories for our simulation cohort participants selected from ADNI. Scores are tracked over number of years elapsed since a participant’s baseline ADNI observation. A) Real individual trajectories for all simulation cohort participants, with overlaid female- and male-specific group-level trends estimated by Eq. 1 indicated in red and blue, respectively, with confidence intervals. These two trends nearly overlap for the 0-to-18-month period that corresponds to the CLARITY AD trial duration. There is no statistically significant sex difference in estimated cognitive decline rate, as the confidence intervals on the slope values overlap between the two models (see Table 2). The models describing these trends were used in generating the Experiment 1 simulations. B) Real individual trajectories for all simulation cohort participants with the cohort-level trend line estimated by Eq. 1 indicated in green, with confidence interval. The model describing this trend was used in generating the Experiment 2 simulations.

**Table 2.** Key estimated parameters from the Eq. 1 model fit to data from A) only male participants, B) only female participants, and C) male and female participants together in the simulation cohort.

| **A) Male-only model**<br>Nakagawa conditional<br>$R^2 = 0.931$ | $\beta$ | Standard<br>error | $t$-value | $p$-value |
| --- | --- | --- | --- | --- |
| Median-centred baseline CDRSB | -0.08145 | 0.03508 | -2.322 | 0.020350 |
| Baseline AD diagnosis status | 0.33407 | 0.11964 | 2.792 | 0.005285 |
| Years since baseline | 0.77923 | 0.06486 | 12.014 | $< 2e-16$ |
| APOE4 status | -0.18959 | 0.04935 | -3.841 | 0.000127 |
| Interaction of median-centred baseline CDRSB with Years since baseline | 0.31434 | 0.04464 | 7.041 | 7.6e-12 |
| **B) Female-only model**<br>Nakagawa conditional<br>$R^2 = 0.928$ | $\beta$ | Standard<br>error | $t$-value | $p$-value |
| Median-centred baseline CDRSB | -0.16455 | 0.04115 | -3.999 | 6.76e-05 |
| Baseline AD diagnosis status | 0.53409 | 0.14458 | 3.694 | 0.00023 |
| Years since baseline | 0.84646 | 0.08339 | 10.151 | $< 2e-16$ |
| APOE4 status | -0.32775 | 0.05849 | -5.604 | 2.64e-08 |
| Interaction of median-centred baseline CDRSB with Years since baseline | 0.35413 | 0.05168 | 6.852 | 4.70e-11 |
| **C) Male and female model**<br>Nakagawa conditional<br>$R^2 = 0.930$ | $\beta$ | Standard<br>error | $t$-value | $p$-value |
| Median-centred baseline CDRSB | -0.11808 | 0.02668 | -4.426 | 9.96e-06 |
| Baseline AD diagnosis status | 0.41625 | 0.09223 | 4.513 | 6.62e-06 |
| Years since baseline | 0.80663 | 0.05113 | 15.777 | $< 2e-16$ |
| APOE4 status | -0.24694 | 0.03774 | -6.543 | 7.12e-11 |
| Interaction of median-centred baseline CDRSB with Years since baseline | 0.33388 | 0.03365 | 9.923 | $< 2e-16$ |

#### 3.2.2. Part 2: Sex-specific trial simulation results

Figure 3 shows histograms of simulated CDRSBΔbl slope differences (i.e., differences in cognitive decline rate) between drug and placebo groups, taken from within 10,000 male-only and 10,000 female-only simulated trials adhering to the CLARITY AD trial design parameters. The horizontal axis of the histogram signifies values of the difference in cognitive decline rate between a trial’s drug and placebo groups. Each value is obtained from within one simulated trial instance and equals the drug-treated group’s slope on its CDRSBΔbl trend minus the placebo group’s slope on its CDRSBΔbl trend. The female-only simulations and the male-only simulations each have their own histogram (red and blue, respectively, in Fig. 3). Each histogram thus illustrates the distribution of observed drug effect sizes across the corresponding 10,000 sex-specific simulations.

**Figure 3.**
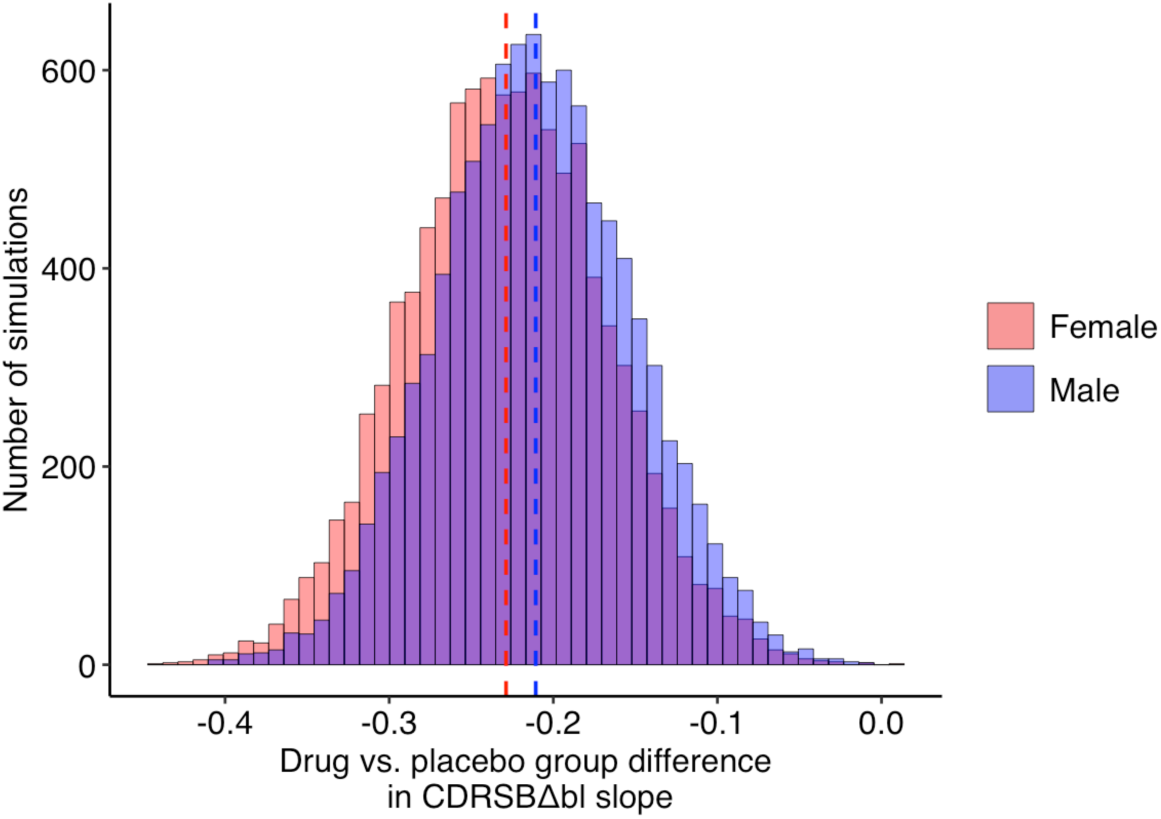
Histograms of simulated CDRSBΔbl slope differences for drug-treated vs. placebo groups (i.e., observed drug effect sizes) for simulated trials adhering to the CLARITY AD design parameters, but including only males (blue) or only females (red). Each histogram represents 10,000 simulations. The mean value for each distribution is indicated by a color-coded dashed line. The small difference in means is statistically significant, as is the difference in variances.

Every simulated trial had the same cohort-level drug effect encoded as a 27% reduction in CDRSBΔbl slope for drug vs. placebo participants. However, the decline rate for the placebo group differed between the female-only and male-only simulations. Within each trial, the placebo decline rate was defined as the *β* for the “*Years*_*since*_*baseline*” term in Table 2; the male-only and female-only models each have their own value for this *β*. Thus, as the drug effect in each trial was encoded as a percent reduction in the placebo decline rate, the 10,000 male-only and 10,000 female-only simulated trials had a different value on average for the observed drug effect. This on-average difference explains the offset positioning of the histograms in Fig. 3.

A Bartlett test of homogeneity of variances showed that the distributions’ variances were significantly different (*K*^2^ = 7.4819, *p* = 0.006232).^43,44^ A two-sample *t*-test showed a statistically significant difference in means for the distributions (*t* = 21.306, *p* < 2.2e−16). The mean effect sizes from the male and female simulations are indicated in Fig. 3. The means are *μ*_male_ = −0.211 and *μ*_female_ = −0.229, and the standard deviations are *σ*_male_ = 0.059 and *σ*_female_ = 0.061. The mean from the male-only simulations is approximately 7.9% less than the mean from the female-only simulations.

This difference is means is linked directly to the statistically nonsignificant sex difference in cognitive decline rate in our ADNI simulation cohort, i.e., the difference between the *β* values for “*Years*_*since*_*baseline*” in Table 2-A and B. This sex difference leads to a corresponding difference in the mean observed effect sizes in our simulations.

When using only 1,000 simulations of the male or female trial scenarios, the statistically significant difference in means persisted (*t* = 5.8396, *p* = 6.097e-09), but the *t-*value decreased and the *p*-value increased compared to using 10,000 simulations. The difference in statistical significance for 1,000 vs. 10,000 simulations is linked primarily to statistical power: The more sex-specific trials we simulate (where there is a true difference in mean drug effect between the male and female simulations), the higher the probability of obtaining a statistically significant estimate for that difference in means. The implication here is that even with a reduced number of sex-specific trials simulated – 1,000 vs. 10,000 – we still obtained a statistically significant estimate for the difference in mean observed drug effect size between the male-only and female-only simulations. Thus, the statistically nonsignificant sex difference in natural cognitive decline rate (summarized in Table 2-A and B) was large enough to produce a sex difference in the mean simulated drug effect when using only 1,000 simulations, a scenario where statistical power to detect the difference in means was much lower compared to using 10,000 simulations.

### 3.3. Experiment 2 results

Figure 4 shows a histogram of 10,000 simulated CDRSBΔbl slope difference values, each signifying the difference in drug effect size between two subgroups of 900 randomly selected participants from within one simulated trial. Male and female participants were included at random in each simulation. The Eq. 1 model, here fit to data from all our simulation cohort participants, was used to generate the simulations. Table 2-C lists key model parameters, where a Nakagawa conditional *R*^2^ of 0.930 indicates a well-fitting model.

**Figure 4.**
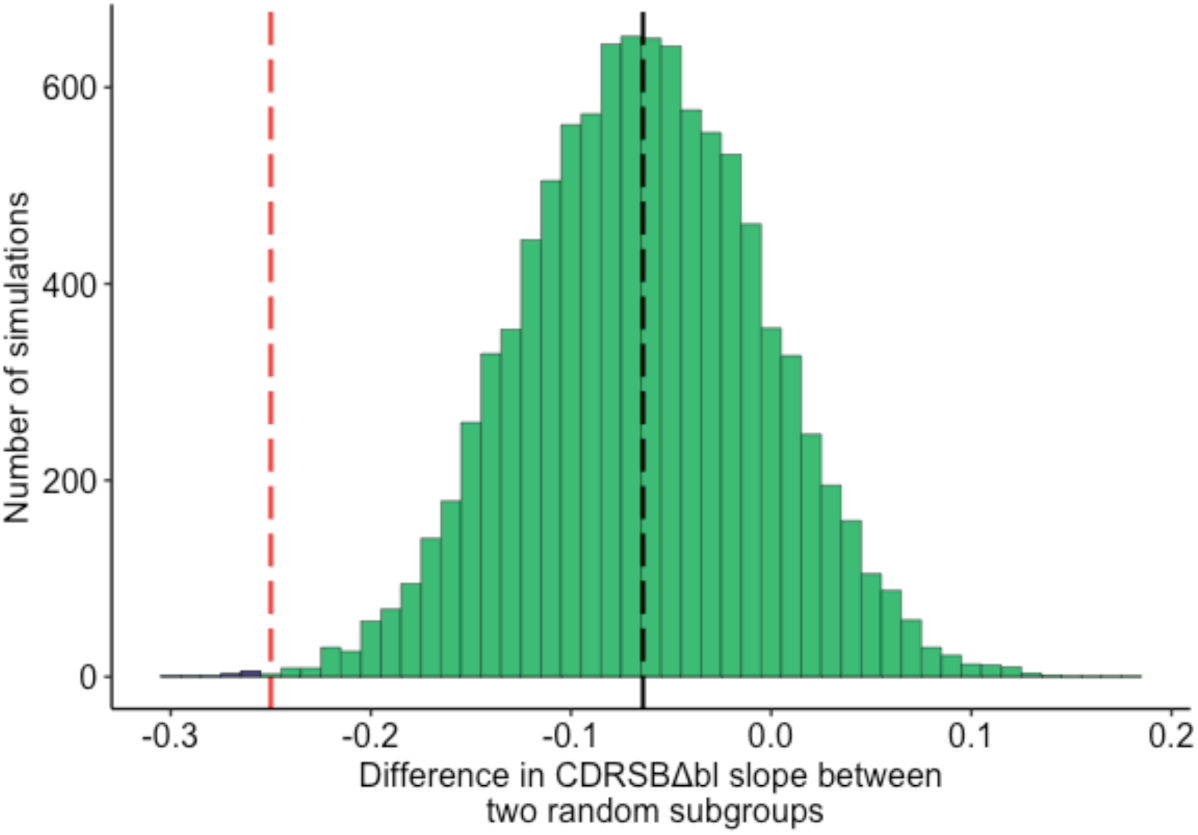
Histogram of differences in CDRSBΔbl slope (here signifying differences in drug effect size) between two subgroups of randomly selected participants in 10,000 trial simulations constrained by the CLARITY AD design. The red dashed line indicates the value corresponding to the 31% difference between sex subgroups reported in CLARITY AD. The black dashed line corresponds to the preloaded known sex difference in drug effect from Experiment 1. Only 12 of 10,000 simulations (indicated in purple to the left of the red line) had a subgroup difference ≥31%. Even when a large difference is preloaded between subgroups, there is an extremely low probability of randomly observing a ≥31% difference in drug effect between subgroups.

The red vertical line in Fig. 4 indicates the difference in drug effect size between subgroups that corresponds to the CLARITY AD-reported 31% difference between its male and female subgroups. A ≥31% difference here implies that one subgroup’s participants have at least a 31% greater reduction in cognitive decline rate compared to the other subgroup.

Additionally, in each simulation we encoded a 7.9% between-subgroup difference in drug effect. With this bias, one subgroup always declines 7.9% slower than the other, resulting in a corresponding on-average difference in decline rate between subgroups across our 10,000 simulations (indicated by the black line in Fig. 4). This 7.9% equals the percent difference in mean drug effect size between males and females in our Experiment 1. This bias effectively preloads the known sex difference in drug effect and thus biases the simulation in favour of CLARITY AD. Any further difference between subgroups is attributable to participant heterogeneity and randomization.

Only 12 of the 10,000 simulated trials had a subgroup difference in drug effect ≥31%, even with our known sex difference in drug effect preloaded. This result signifies a 0.0012 probability of observing a drug effect difference ≥31% between two subgroups, even when there is a known difference of 7.9% between the subgroups.

## 4. Discussion

We examined whether the sex difference in lecanemab’s clinical effect observed in CLARITY AD could be explained by inherent sex differences in cognitive decline, or as a fluke linked to participant heterogeneity and randomization. While our selection of ADNI participants who met CLARITY AD’s inclusion and exclusion criteria resulted in a female cohort that was slightly younger and less educated (age: 71.6 ± 7.3 years, education: 15.3 ± 2.7 years) than the males (age: 73.9 ± 6.8 years, education: 16.4 ± 2.8 years), the groups had similar proportions of APOE4 positive individuals and a similar split of MCI and mild AD participants.

The yearly change in CDRSB, estimated in Experiment 1 Part 1 from the *β* values of the fitted sex-specific models (Eq. 1), was not statistically significantly different between males (*β*_male_ = 0.77923 ± 0.06486) and females (*β*_female_ = 0.84646 ± 0.08339). In other words, even though the graph in Fig. 2-A appears to show a slight sex-related difference, the confidence intervals (in grey) overlap for the 10+ year period fitted, and the two regression lines overlap almost completely in the 0-to-18-month period that corresponds to the CLARITY AD trial duration.

Nonetheless, our simulation results from Experiment 1 Part 2 showed a statistically significant difference in mean drug effect size between male-only and female-only simulated trials constrained by the CLARITY AD design parameters. Males had a mean observed effect that was approximately 7.9% smaller than in females. However, that difference does not explain the 31% difference in drug effect between males and females in CLARITY AD. Moreover, CLARITY AD reported a larger effect for males than females, while we found a smaller effect for males.

Based on these results, the answer to our Experiment 1 question is: No, the CLARITY AD sex difference cannot be explained by lecanemab-independent sex differences in cognitive decline. Our simulations provide an empirical justification for this conclusion.

Experiment 2 showed that a ≥31% difference in drug effect between subgroups is extremely unlikely to occur as a fluke, even when a known difference between subgroups is present. Based on our empirical simulations, the estimated probability of that occurrence is 0.0012. Experiment 2 produces the same conclusion as an expert reading of the key CLARITY AD forest plot (Fig. S1-B in the cited paper’s appendix), where the confidence intervals on the male and female mean effects do not overlap the mean effects of the opposing sex.^7^ This non-overlap indicates a statistically significant sex difference in lecanemab’s effect. The answer to our Experiment 2 question is thus: No, the CLARITY AD sex difference cannot be explained as a fluke difference between the male and female subgroups.

Overall, these results imply that the magnitude of the CLARITY AD sex difference is unlikely to be explained by the phenomena we investigated. CLARITY AD also showed statistically significant larger effects for lecanemab in males than in females in two of three non-primary clinical endpoints (ADCOMS and ADCS-MCI-ADL).^7^ The point estimate of lecanemab’s effect was numerically larger in males than females for the third non-primary endpoint, ADAS-Cog14.^7^ However, the sex subgroup analysis of four quality-of-life (QOL) metrics showed no statistically significant sex difference in QOL change since baseline at 18 months.^47^ Nonetheless, point estimates on three of the metrics (Zarit Burden Interview total score and QOL in AD total score [A] by subject and [B] by subject by proxy) trended toward larger benefit in males than females.^48,49^

Given these outcomes, the CLARITY AD result likely represents a genuine sex difference in lecanemab’s clinical effect. Further research is needed to determine whether such a difference could be related to biological or gender differences, such as in hormone profiles, cognitive reserve, and education.^2^ More generally, our study points to a possible sex difference in the clinical efficacy of amyloid-targeting drugs, specifically for lecanemab, and possibly for those with different mechanisms of action such as aducanumab and donanemab.^6–8^

In aducanumab’s Phase 3 trial EMERGE, a statistically significant primary clinical effect was reported only for the male subgroup in the main results paper’s supplementary Fig. 3-A forest plot.^6^ However, there was no significant sex difference in that effect. Like lecanemab, aducanumab trended toward effectiveness in females, and the nonsignificance of the effect could be explained by low power. Similarly, no significant sex difference in primary efficacy was observed in donanemab’s positive Phase 3 trial TRAILBLAZER-ALZ 2.^8^ However, the eFigure 9-B forest plot in the results paper’s supplement indicates a statistically nonsignificant effect in males only. This nonsignificance could again be due to low power.

These outcomes suggest that CLARITY AD’s sex difference could be linked to lecanemab’s mechanism of action. Unfortunately, to the best of our knowledge, sex-specific data on amyloid clearance are not publicly available for the trials discussed. For CLARITY AD, sex subgroup analyses of amyloid PET data might reveal whether lecanemab’s sex difference in efficacy is accompanied by corresponding sex differences in amyloid clearing. Potential differences in the clinical impact of removing amyloid would fit conceptually with previously observed modulating effects of sex on the association between AD pathology and clinical progression. For example, a 2005 study of 141 older Catholic clergy members showed that amyloid plaques and neurofibrillary tangles were more likely to manifest clinically as dementia in females than in males.^25^ On the other hand, the presence of an amyloid clearance sex difference could explain the observed sex difference in lecanemab’s clinical efficacy. A comparison of CLARITY AD’s sex disaggregated PET data with corresponding data from the Phase 3 aducanumab and donanemab trials could highlight possible links between a drug’s action mechanism and any sex differences in amyloid clearing and clinical efficacy.

While CLARITY AD had near-equal sex representation (51.6% and 53.0% female in lecanemab and placebo groups, respectively), future trials of amyloid-targeting drugs might benefit from recruiting larger cohorts overall while preserving sex parity. Larger sample sizes could ensure adequate power to more definitively observe sex differences, including in preclinical trial phases and drug development research.^22^ Stratification within sex subgroups, such as by menopausal state or gender-biased risk factors (e.g., lower education in women than men),^50,51^ might help disentangle sex and gender contributions to a trial’s potentially observed male/female difference in a drug’s clinical effect.

In addition to the lecanemab sex difference addressed in the present study, CLARITY AD reported differences in observed drug effect for older vs. younger participants, and between APOE4 non-carrier, homozygotes, and heterozygotes.^7^ These outcomes suggest that detailed within-subgroup analyses are needed for demographic categories in addition to sex. Future trials would benefit from larger cohorts that enable high powered subgroup analyses generally, and potentially within subgroups. Such analyses could enable more precise definitions of drug indications and more personalized patient care. Fortunately, novel trial enrichment strategies (e.g., using cognitive decline prediction tools) could enhance a trial’s power to detect effects within subgroups without requiring an increase in the total number of participants.^52,53^ Advanced trial designs such as those relying on digital twins might also require fewer participants than conventional randomized placebo-controlled trials in AD.^54^ To date, however, such tools are not widely used.

Low powered subgroup analyses in contemporary AD trials should be reported and interpreted cautiously. As evidenced by recent works citing the CLARITY AD sex difference, the reporting of AD trial subgroup results in forest plots might lead to misinterpretations of statistically nonsignificant effects in low powered subgroups. In response to criticism that lecanemab might not benefit females, the CLARITY AD paper’s authors themselves stated that the trial was not powered to evaluate subgroups separately.^19^ The Phase 3 aducanumab and donanemab trials described earlier could also suffer from such misinterpretations. Unless a trial is powered to evaluate efficacy within each subgroup, single *p*-values (and/or confidence intervals) in subgroup analyses should only be used to indicate whether a treatment was significantly more effective in one subgroup than another. AD trials should still be explicitly powered to evaluate such differences.

Overall, our analysis has some limitations. We used a linear modeling technique to generate our simulations, but AD is known to progress nonlinearly over the full disease timeline.^55^ However, our simulations cover trajectories over 18 months only, i.e., the CLARITY AD trial duration. Linear assumptions may be justifiable here. Also, linear mixed effects models can capture inter-participant heterogeneity in cognitive decline rate around a cohort-level average trend, even across participants with MCI or mild dementia.^56^ Future analyses might benefit from nonlinear models, including ones that make no assumptions about the shape of disease trajectories when extracting drug effects from trial data.^56^

We also do not include genuine placebo effects in our modeling, so our simulations essentially compare a drug effect to the ADNI standard of care. ADNI participants were recruited across Canada and the United States, where care standards can differ between and within the countries.^57,58^ The demographic profile of ADNI also does not represent the diversity in CLARITY AD. ADNI participants are predominantly highly educated, white, and male, while CLARITY AD recruited from many sites across three large geographic regions (North America, Europe, and Asia-Pacific).^7^ Given ADNI’s limited scope, we did not account for geography in our simulations. Future work might use publicly available data from past international trials to do so (e.g., from the Critical Path for Alzheimer’s Disease).^59^

To conclude, our simulation results combine with the published CLARITY AD sex outcome to suggest that lecanemab might have lower clinical benefit in females than in males. This result has direct implications for AD treatment decisions in the clinic, drug approval decisions in regulatory agencies internationally, and design considerations for future trials of amyloid-targeting drugs for AD. Results from CLARITY AD’s open-label extension might help clarify the longer-term impact of lecanemab in females.

The FDA has approved three anti-amyloid drugs since 2021: aducanumab, lecanemab, and donanemab.^14,60,61^ Considering CLARITY AD’s results, certain types of amyloid-targeting drugs might function differently in females and males. Research into possible mechanisms could be accelerated by drug developers allowing researchers secure access to patient data from trials like lecanemab’s CLARITY AD,^7^ donanemab’s TRAILBLAZER-ALZ 2,^8^ aducanumab’s EMERGE/ENGAGE,^6^ and others.

## Data Availability

Data used in preparation of this article were obtained from the Alzheimer's Disease Neuroimaging Initiative (ADNI) database (https://adni.loni.usc.edu).

## 6. Acknowledgments

Data collection and sharing for this project was funded by the Alzheimer’s Disease Neuroimaging Initiative (ADNI) (National Institutes of Health Grant U01 AG024904) and DOD ADNI (Department of Defense award number W81XWH-12-2-0012). ADNI is funded by the National Institute on Aging, the National Institute of Biomedical Imaging and Bioengineering, and through generous contributions from the following: AbbVie, Alzheimer’s Association; Alzheimer’s Drug Discovery Foundation; Araclon Biotech; BioClinica, Inc.; Biogen; Bristol-Myers Squibb Company; CereSpir, Inc.; Cogstate; Eisai Inc.; Elan Pharmaceuticals, Inc.; Eli Lilly and Company; EuroImmun; F. Hoffmann-La Roche Ltd and its affiliated company Genentech, Inc.; Fujirebio; GE Healthcare; IXICO Ltd.; Janssen Alzheimer Immunotherapy Research & Development, LLC.; Johnson & Johnson Pharmaceutical Research & Development LLC.; Lumosity; Lundbeck; Merck & Co., Inc.; Meso Scale Diagnostics, LLC.; NeuroRx Research; Neurotrack Technologies; Novartis Pharmaceuticals Corporation; Pfizer Inc.; Piramal Imaging; Servier; Takeda Pharmaceutical Company; and Transition Therapeutics. The Canadian Institutes of Health Research is providing funds to support ADNI clinical sites in Canada. Private sector contributions are facilitated by the Foundation for the National Institutes of Health (www.fnih.org). The grantee organization is the Northern California Institute for Research and Education, and the study is coordinated by the Alzheimer’s Therapeutic Research Institute at the University of Southern California. ADNI data are disseminated by the Laboratory for Neuro Imaging at the University of Southern California.

The present study was supported by a donation from the Famille Louise & André Charron and a research grant (Principal Investigator: D. Louis Collins) from the Canadian Institutes of Health Research (CIHR). Daniel Andrews is supported by McGill University and by a doctoral research scholarship from CIHR. Dr. Louis Collins reports receiving research funding from CIHR, the Natural Sciences and Engineering Research Council of Canada, Brain Canada, the Weston Foundation, and the Famille Louise & André Charron.

## 7. Consent statement

ADNI received ethics approval from all participating institutions and received informed from all participants or their study partner.

## 8. Conflict of interest statement

Daniel Andrews declares no conflict of interest. Dr. Simon Ducharme has conducted sponsored research with the following companies in the last two years: Biogen, Novo Norkisk, Janssen, Innodem Neurosciences and Alnylam. Dr. Ducharme has participated in advisory committees or received speaker fees from the following companies in the last two years: Eisai, QuRALIS, Eli Lilly, NKGen, and IntelGenX. Dr. Chertkow has been supported by a Foundation Grant from the CIHR (Canadian Institutes for Health Research), along with funding from the National Institutes of Health (USA), the Weston Foundation and the Baycrest Health Sciences Foundation. Dr. Chertkow has participated as a site PI in pharmaceutical trial activities in the past five years sponsored by: Hoffmann-La Roche Limited, TauRx, Lilly, Anavex Life Sciences, Alector LLC, Biogen, Esai, and Immunocal (site investigator for trials). Dr. Chertkow has participated as an unpaid advisor in 2020 for establishment of an international database by Biogen. Dr. Chertkow has participated in advisory boards for Esai and Lilly Co., with honoraria going to the Rotman Research Institute. Dr. Chertkow is Scientific Director for the Canadian Consortium on Neurodegeneration in Aging (CCNA), which receives partner support from a set of partners including: Industry: Pfizer Inc., Lilly, and Sanofi; Not for profit organizations: Brain Canada, Alzheimer Society of Canada, Women’s Brain Health Initiative, Picov Family Foundation, New Brunswick Health Research Foundation, Saskatchewan Health Research Foundation, and the Ontario Brain Institute. Dr. D. Louis Collins holds an equity interest in True Positive Medical Devices Inc.

## Notes

### Author Declarations

Data used in preparation of this article were obtained from the Alzheimer's Disease Neuroimaging Initiative (ADNI) database (https://adni.loni.usc.edu) after our data request application was approved by ADNI (https://adni.loni.usc.edu/data-samples/access-data/). ADNI received ethics approval from all participating institutions and received informed consent from all participants.

